# Development and validation of nomogram to predict long-term prognosis of critically ill patients with acute myocardial infarction

**DOI:** 10.1101/2020.08.14.20174953

**Authors:** Yiyang Tang, Lihuang Zha, Xiaofang Zeng, Yilu Feng, Wenchao Lin, Zhenghui Liu, Zaixin Yu

## Abstract

**Background:** Acute myocardial infarction (AMI) is a common critical illness in the cardiovascular field, with poor prognosis. This study aimed to construct a nomogram to predict long-term survival of critically ill patients with AMI, which helps to assess severity, guide treatment, and improve prognosis.

**Methods and results:** The clinical data of patients with AMI was extracted from the database MIMIC-III v1.4. The Cox proportional hazards models were performed to identify the independently prognostic factors, and a nomogram for predicting long-term survival of AMI patients was developed based on the multifactor analysis, of which discriminative ability and accuracy was evaluated by concordance index (C-index) and calibration curve.

**Results:** A total of 1202 patients were included in the analysis, of which 841 were divided into the training set and 361 were the validation. Multivariate analysis shown that age, blood urea nitrogen, respiratory rate, SAPSII score, cardiogenic shock, cardiac arrhythmias, and respiratory failure served as the independently predictive factors, which were incorporated into the nomogram. Moreover, the nomogram shown favorable performance for predicting 4-year survival of AMI patients with the C-index of 0.788 [95% confidence interval (CI): 0.763 to 0.813] and 0.783 (95% CI: 0.748 to 0.818) in the training and validation set, respectively.

**Conclusion:** The nomogram we constructed here can accurately predict the long-term survival of patients with AMI.

## 1 Introduction

Acute myocardial infarction (AMI) is defined as death or necrosis of myocardial cell caused by acute, severe, and sustained ischemia and hypoxia after coronary artery occlusion, and it is the most serious subtype of coronary heart disease [1]. The overall prevalence of AMI among adults over the age of 20 years old in the United States is 3.0% according to NHANES data from 2013 to 2016, and an American will suffer from AMI approximately every 40 seconds [2]. Moreover, the prognosis of AMI is poor, with a 5-year mortality rate as high as 51%, which heavily threatens human health and causes a great socioeconomic burden [3]. In view of this, the identification and development of risk factors or models for identifying high-risk patients is of great significance to improve the prognosis of patients, because advanced interventions can be timely taken.

There have been several risk scores constructed to predict the prognosis of patients with AMI, among which the Thrombolysis in Myocardial Infarction (TIMI) [4] and the Global Registry of Acute Coronary Events (GRACE) [5] scores are the most widely used. Regrettably, these scores are mainly suitable for the short-term prognosis evaluation such as in-hospital mortality and long-term survival prediction based on these risk scores used for short-term prognosis prediction is flawed in accuracy [6]. With the great progress made in the diagnosis and treatment of AMI in the last few decades, especially the widespread adoption of emergency percutaneous coronary intervention, the survival time of patients with AMI has been significantly prolonged and it has become urgent to establish a prognostic model to predict the long-term outcomes of patients [7].

Nomogram, a risk score tool for medical decision-making, has the characteristics of simple operation and easy understanding by visualizing the results of the prediction model [8]. In clinical practice, the total score of patients can be calculated according to the respective score of each predictor in the nomogram, and then the probability of the specific disease-related outcome can be obtained, which has been successfully applied to various diseases including septic acute kidney injury [9], acute type A aortic dissection [10], and heart failure [11]. In the present study, we used the clinical records from the public database to conduct and validate a nomogram to predict the long-term overall survival of critically ill patients with AMI.

## 2 Materials and Methods

### 2.1 Data source

The clinical data for analysis was downloaded from the freely available critical care database, the Multiparameter Intelligent Monitoring in Intensive Care III version 1.4 (MIMIC-III v1.4) [12], which is run by the Massachusetts Institute of Technology and funded by the National Institutes of Health (NIH). The database recorded detailed information from over forty thousand de-identified patients in Beth Israel Deaconess Medical Center between 2001 and 2012, including demographic data, vital signs, comorbidities, and laboratory tests, which provides reliable data resource for clinicians to conduct epidemiological studies. Besides, the database provides the patient’s death time inside and outside the hospital from the hospital database or the social security database. The date of death outside the hospital is stored in two systems, namely the CareVue system with four years follow-up and the MetaVision with 90-days. Before implementing this research, author Tang completed and passed the CITI “Data or Specimens Only Research” course (No.9014457) and obtained authorization for database access. In need of special note here is that this database was approved by the institutional review boards of Massachusetts Institute of Technology (Cambridge, MA, USA) and Beth Israel Deaconess Medical Center (Boston, MA, USA), and no additional ethical approval need to be provided here.

### 2.2 Statement

According to the requirements of the database, author Tang has completed the required training course, CITI “Data or Specimens Only Research” course, and passed the corresponding exams (record ID: 35980937) to get the access permission. Since this project has been approved by the institutional review boards of Massachusetts Institute of Technology (Cambridge, MA, USA) and Beth Israel Deaconess Medical Center (Boston, MA, USA) and the identifying elements related to patient privacy have been removed from the database, our research does not need to provide the additional approval of ethics committee.

### 2.3 Participants and Design

The process of inclusion and exclusion of participants was presented in Figure 1. Among the 46,520 patients in the MIMIC-III database, patients first admitted to the intensive care unit (ICU) and diagnosed with AMI based on the ninth revision of the international classification of diseases code (ICD-9) were included in our research. Patients under the age of 18 with ICU stay less than 24 hours and survival time less than 0 were excluded from our study. Besides, patients in the MetaVision system are also excluded due to the shorter follow-up time as mentioned earlier [13]. Randomly select 70% of the eligible patients as the training set to establish the prediction model, and use the remaining 30% of the patients as the validation set to verify the prediction performance of the model. The primary clinical endpoint of our research was overall survival (OS), defined as the time from ICU admission to death or the last date of follow-up (4 years).

**Figure 1.**
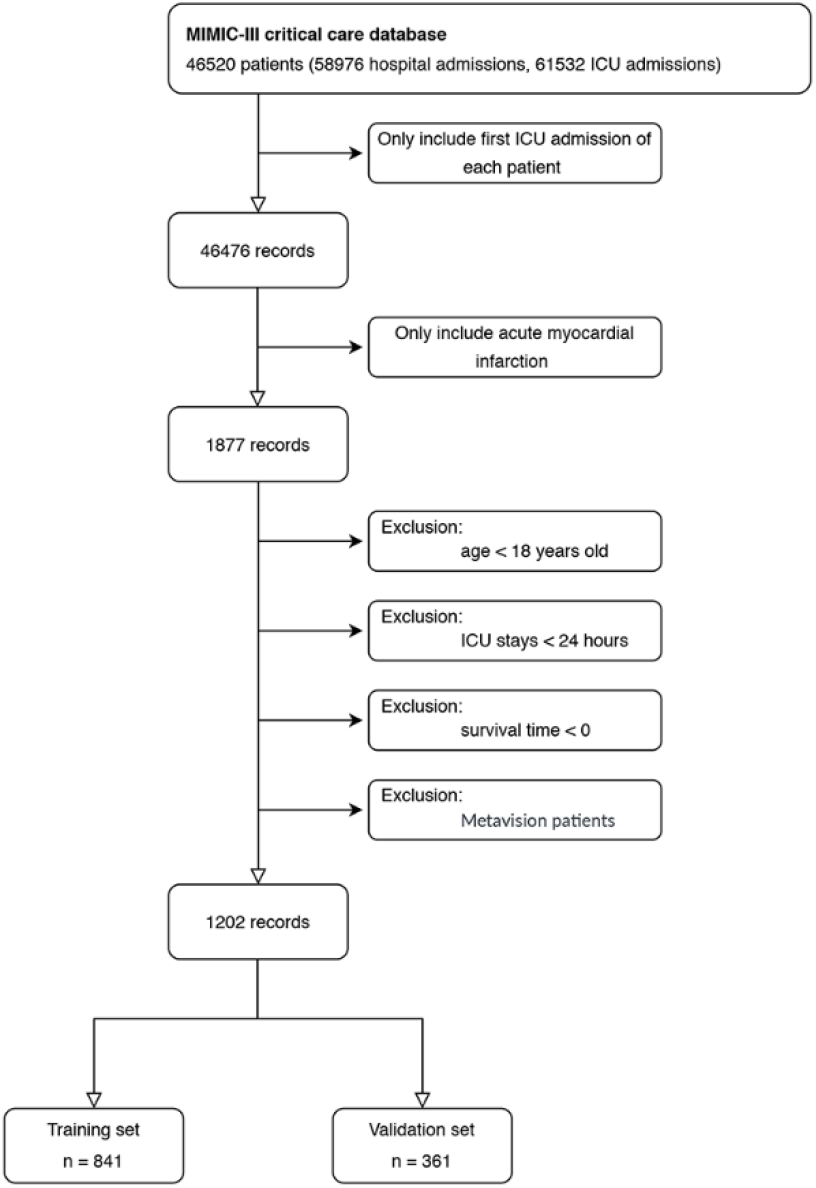
Workflow of the inclusion and exclusion of the study subjects. ICU, intensive care unit.

### 2.4 Data Extraction

The clinical data of each patients, including demographic parameters, vital signs, comorbidities, laboratory test, scoring systems, and interventions, was extracted from MIMIC-III database with Structured Query Language in PostgreSQL tools (version 9.6). Demographic parameters mainly referred to age, gender and ethnicity (White, Black, and others), while vital signs included body temperature, systolic blood pressure (SBP), diastolic blood pressure (DBP), heart rate (HR), respiratory rate (RR), and percutaneous oxygen saturation (SpO2). It is worth noting that the age of patients over 89 years old has been fixed to be 300 for the purpose of protecting patient privacy and we have converted the age of these patients (real age = age - 300 +89) [14]. Comorbidities were also extracted with corresponding ICD-9 code, including cardiogenic shock, cardiac arrest, congestive heart failure, cardiac arrhythmias, valvular heart diseases, pulmonary circulation diseases, peripheral vascular diseases, hypertension, diabetes, pneumonia, respiratory failure, liver disease, renal failure, stroke, depression, and hypothyroidism. We used the Sequential Organ Failure Assessment score (SOFA) [15], the Simplified Acute Physiology Score II (SAPSII) [16], and the Glasgow Coma Scale score (GCS) [17, 18] to assess the severity of patients, which were calculated according to the physiological and laboratory parameters in admission. Laboratory tests included white blood cells (WBC), hemoglobin, platelets, anion gap, sodium, potassium, chloride, bicarbonate, creatinine, blood urea nitrogen (BUN), glucose, prothrombin time (PT), activated partial thromboplastin time (APTT), troponin T (cTnT), and lactate within the first 24 hours after ICU admission. Interventions contained the use of vasopressor medicine, dialysis, mechanical ventilation and percutaneous coronary intervention (PCI). Except for cTnT and lactate, the missing values of the other variables are all within 10%, which were filled by multivariate multiple imputation with chained equations while cTnT and lactate were regarded as dummy variables for following statistical analysis to reduce the possible bias of simple filling [19].

### 2.5 Statistical Analysis

The continuous variables were presented in the form of mean ± standard deviation (SD) with Kruskal Wallis test for hypothesis test. The categorical variables are expressed as numbers (percentages), which were analyzed by using Chi-square or Fisher’s exact tests, as appropriate.

The clinical data of training set was used to construct the nomogram. First, we conducted the univariate Cox regression analysis to explore possible variables that may be related to the OS of patients with AMI. Then, the multivariable Cox regression analysis with forward stepwise selection was performed on these significant variables in univariate analyses (P < 0.05). Last, we established the nomogram to visualize the results of multivariate analysis by using the rms R package. And variance inflation factor (VIF) was calculated to test the collinearity between variables, with 2 as the threshold.

In validation set, the risk scores of each patient based on the result of training set was regarded as a variable to conduct Cox proportional hazard regression. The concordance index (C-index) was calculated with Hmisc R package to assess the discrimination of the model for prognosis. The calibration curve with bootstrap method and 1000 resample was used to reflect the consistency between the actual probability and predicted by the nomogram. The sensitivity and specificity of the nomogram were evaluated by the area under the curve (AUC) value of the receiver operating characteristic (ROC) curve with survivalROC R package.

All statistical analyses of our study were implemented through Stata16.0 (StataCorp LLC, College Station, Tex) and R software version 3.5.3 (http://www.r-project.org/). P < 0.05 (2-sided) was considered statistically significant.

## 3 Results

### 3.1 Baseline characteristics of Subjects

After screening the study subjects according to the inclusion and exclusion criteria, a total of 1202 patients with AMI were included in the study, of which 841 patients were randomly entered onto the training set, and 361 patients were analyzed as the validation set. Overall, most of patients are male (770, 64.1%) and White (736, 61.2%), and the age of the subjects was generally older, with a median of 68.9. The detailed clinical characteristics of the patients in the training and validation were shown in Table 1. There were no significant differences in most variables between the two groups. The 4-year OS rates were 65.9%, while in training and validation set were 66.8% and 63.7%, respectively.

**Table 1.** The characteristics of critical ill patients with AMI in the training and validation sets.

| Variable | Cohort |  | P-value |
| --- | --- | --- | --- |
|  | Training (n = 841) | Validation (n = 361) |  |
| Age (years) | 68.2 ± 13.7 | 67.4 ± 14.3 | 0.477 |
| Gender, n (%) |  |  | 0.922 |
| Female | 303 (36.0) | 127 (35.7) |  |
| Male | 538 (64.0) | 232 (64.3) |  |
| Ethnicity, n (%) |  |  | 0.468 |
| White | 512 (60.9) | 224 (62.0) |  |
| Black | 30 (3.6) | 8 (2.2) |  |
| Other | 299 (35.6) | 129 (35.7) |  |
| HR, beats/minute | 83.2 ± 15.0 | 82.9 ± 16.0 | 0.678 |
| RR, beats/minute | 18.4 ± 3.6 | 18.4 ± 3.4 | 0.845 |
| Temperature, °C | 36.9 ± 0.6 | 36.9 ± 0.6 | 0.878 |
| SBP, mmHg | 112.4 ± 15.0 | 112.3 ± 14.6 | 0.987 |
| DBP, mmHg | 59.9 ± 9.5 | 59.9 ± 8.9 | 0.594 |
| SpO<sub>2</sub>, % | 97.5 ± 1.7 | 97.3 ± 2.2 | 0.234 |
| Weight, Kg | 80.6 ± 19.8 | 80.5 ± 19.2 | 0.880 |
| Scoring systems |  |  |  |
| SOFA | 3.7 ± 3.0 | 3.6 ± 3.1 | 0.431 |
| SAPSII | 35.1 ± 14.2 | 33.9 ± 13.8 | 0.172 |
| GCS | 14.3 ± 2.5 | 14.5 ± 2.0 | 0.652 |
| Dialysis, n (%) | 45 (5.4) | 10 (2.8) | 0.050 |
| Vasopressor, n (%) | 139 (16.5) | 65 (18.0) | 0.532 |
| Ventilation, n (%) | 229 (27.2) | 97 (26.9) | 0.898 |
| PCI, n (%) | 474 (56.4) | 207 (57.3) | 0.753 |
| 4-year overall survival | 562 (66.8) | 230 (63.7) | 0.297 |
|  | Training (n = 841) | Validation (n = 361) |  |
| Comorbidities, n (%) |  |  |  |
| Cardiogenic shock | 152 (18.1) | 51 (14.1) | 0.094 |
| Cardiac arrest | 63 (7.5) | 29 (8.0) | 0.746 |
| CHF | 70 (8.3) | 25 (6.9) | 0.410 |
| Cardiac arrhythmias | 50 (5.9) | 16 (4.4) | 0.291 |
| Pulmonary circulation | 5 (0.6) | 2 (0.6) | 1.000 |
| Peripheral vascular | 62 (7.4) | 29 (8.0) | 0.691 |
| Valvular heart disease | 25 (3.0) | 7 (1.9) | 0.308 |
| Hypertension | 65 (7.7) | 14 (3.9) | 0.014 |
| Diabetes | 223 (26.5) | 68 (18.8) | 0.004 |
| Pneumonia | 116 (13.8) | 40 (11.1) | 0.200 |
| Respiratory failure | 91 (10.8) | 37 (10.2) | 0.769 |
| Liver diseases | 12 (1.4) | 3 (0.8) | 0.394 |
| Renal failure | 87 (10.3) | 23 (6.4) | 0.029 |
| Stroke | 24 (2.9) | 12 (3.3) | 0.661 |
| Depression | 16 (1.9) | 9 (2.5) | 0.511 |
| Hypothyroidism | 19 (2.3) | 25 (6.9) | 0.285 |
| Laboratory test |  |  | 0.050 |
| WBC (K/ul) | 12.9±5.4 | 12.8±5.2 | 0.876 |
| Platelet (K/ul) | 225.1±97.1 | 230.0±91.2 | 0.288 |
| Hemoglobin (g/dl) | 11.7±2.1 | 11.8±2.2 | 0.753 |
| Creatinine (mg/dl) | 1.3±1.2 | 1.1±0.9 | 0.090 |
| BUN (mg/dl) | 24.0±16.5 | 22.7±15.9 | 0.148 |
| Glucose (mg/dl) | 165.2±86.1 | 161.7±74.7 | 0.628 |
|  | Training (n = 841) | Validation (n = 361) |  |
| Laboratory test |  |  |  |
| Sodium (mmol/L) | 138.2±3.9 | 138.1±3.9 | 0.711 |
| Potassium (mmol/L) | 4.2±0.7 | 4.2±0.6 | 0.974 |
| Chloride (mmol/L) | 105.1±5.2 | 104.7±4.9 | 0.307 |
| Bicarbonate (mmol/L) | 22.6±4.1 | 22.9±3.9 | 0.437 |
| PT (second) | 14.4±3.2 | 14.4±3.5 | 0.066 |
| APTT (second) | 53.7±37.9 | 51.9±36.4 | 0.297 |
| cTnT, n (%) |  |  | 0.554 |
| < 1 ng/mL | 654 (77.8) | 271 (75.1) |  |
| ≥ 1 ng/mL | 187 (22.2) | 90 (24.9) |  |
| Lactate, n (%) |  |  | 0.427 |
| <2 mmol/L | 636 (75.6) | 285 (78.9) |  |
| ≥2 mmol/L | 205 (24.4) | 76 (21.1) |  |
| Site of infarction |  |  | 0.471 |
| Anterolateral wall | 59 (7.0) | 24 (6.6) |  |
| Anterior wall | 249 (29.6) | 117 (32.4) |  |
| Inferolateral wall | 38 (4.5) | 19 (5.3) |  |
| Inferoposterior wall | 68 (8.1) | 23 (6.4) |  |
| Inferior wall | 233 (27.7) | 110 (30.5) |  |
| Lateral wall | 23 (2.7) | 4 (1.1) |  |
| Other specified sites | 27 (3.2) | 9 (2.5) |  |
| Unspecified sites | 144 (17.1) | 55 (15.2) |  |

Abbreviations: AMI, acute myocardial infarction; HR, heart rate; RR, respiratory rate; SBP, systolic blood pressure; DBP, diastolic blood pressure; SpO<sub>2</sub>, percutaneous oxygen saturation; SOFA, stroke, and malignancy. Calculate the sequential organ failure assessment score; SAPSII, simplified acute physiology score II; GCS, the Glasgow Coma Scale score. PCI, percutaneous coronary intervention. CHF, congestive heart failure; WBC, white blood cell; BUN, blood urea nitrogen; PT, prothrombin time; APTT, activated partial thromboplastin time; cTnT, troponin T.

### 3.2 Univariate and Multivariate Analysis

The Cox proportional hazard model was applied to identify the prognostic factors of patients with AMI. The variables in the Table 1 were respectively introduced into the univariate Cox regression analysis. A total of 33 variables, such as age, gender, and hemoglobin, served as significant factors for the overall survival of patients with AMI, and the results were presented in Table 2. These variables with P < 0.05 were further included in the multivariate Cox regression analysis. Through the stepwise forward regression method, age, HR, RR, BUN, cardiogenic shock, dialysis, mechanical ventilation seven variable were finally identified as independent prognostic factors for 4-year OS of patients with AMI (hazard ratio: 1.012 to 2.223, P < 0.01, Table 3). The VIF value of all these variables was less than 2, indicating that no linear correlation was observed here.

**Table 2.** Univariate Cox regression analysis of 4-year overall survival in the training set.

| Variable | Univariable analysis |  |  |
| --- | --- | --- | --- |
|  | HR | 95% CI | P-value |
| Age (years) | 1.05 | 1.04 – 1.06 | < 0.0001 |
| Gender, male | 0.65 | 0.51 – 0.82 | 0.0004 |
| GCS | 0.92 | 0.89 – 0.96 | < 0.0001 |
| Heart rate | 1.02 | 1.01 – 1.03 | < 0.0001 |
| Respiratory rate | 1.08 | 1.05 – 1.12 | < 0.0001 |
| Systolic blood pressure | 0.99 | 0.98 – 1.00 | 0.0070 |
| Diastolic blood pressure | 0.96 | 0.95 – 0.97 | < 0.0001 |
| Weight | 0.98 | 0.98 – 0.99 | < 0.0001 |
| Dialysis | 3.68 | 2.58 – 5.26 | < 0.0001 |
| Vasopressor | 1.98 | 1.51 – 2.60 | < 0.0001 |
| Mechanical ventilation | 2.28 | 2.23 – 3.58 | < 0.0001 |
| PCI | 0.60 | 0.48 – 3.58 | < 0.0001 |
| Cardiogenic shock | 2.61 | 2.02 – 3.38 | < 0.0001 |
| CHF | 3.10 | 2.27 – 4.22 | < 0.0001 |
| Valvular heart disease | 1.89 | 1.10 – 3.23 | 0.0205 |
| Pulmonary circulation | 6.38 | 2.37 – 17.17 | 0.0002 |
| Cardiac arrhythmias | 3.38 | 2.39 – 4.78 | < 0.0001 |
| Hypertension | 1.82 | 1.27 – 2.60 | 0.0011 |
| Pneumonia | 2.88 | 2.20 – 3.77 | < 0.0001 |
| Respiratory failure | 3.36 | 2.52 – 4.48 | < 0.0001 |
| Renal failure | 2.18 | 1.60 – 2.97 | < 0.0001 |
| Anterior wall infarction | 0.58 | 0.37 – 0.92 | 0.0190 |
|  | HR | 95% CI | P-value |
| WBC | 1.04 | 1.02 – 1.06 | 0.0005 |
| Hemoglobin | 0.84 | 0.80 – 0.89 | < 0.0001 |
| Anion gap | 1.15 | 1.12 – 1.18 | < 0.0001 |
| Potassium | 1.28 | 1.09 – 1.51 | 0.0033 |
| Bicarbonate | 0.91 | 0.89 – 0.94 | < 0.0001 |
| BUN | 1.03 | 1.03 – 1.04 | < 0.0001 |
| Creatinine | 1.31 | 1.24 – 1.39 | < 0.0001 |
| Glucose | 1.00 | 1.00 – 1.00 | < 0.0001 |
| PT | 1.05 | 1.02 – 1.07 | 0.0011 |
| Lactate ($\geq 2$ mmol/L) | 1.83 | 1.35 – 2.46 | < 0.0001 |

Abbreviations: GCS, the Glasgow Coma Scale score; PCI, percutaneous coronary intervention; CHF, congestive heart failure; WBC, white blood cell; BUN, blood urea nitrogen; PT, prothrombin time; HR, hazard ratio; CI, confidence interval.

**Table 3.** Multivariate Cox regression analysis of 4-year overall survival in the training set.

| Variable | Multivariate analysis |  |  |
| --- | --- | --- | --- |
|  | HR | 95% CI | P-value |
| Age | 1.05 | 1.04 – 1.06 | < 0.0001 |
| Heart rate | 1.01 | 1.00 – 1.02 | 0.0042 |
| Respiratory rate | 1.07 | 1.03 – 1.10 | < 0.0001 |
| Blood urea nitrogen | 1.02 | 1.01 – 1.02 | < 0.0001 |
| Cardiogenic shock | 1.83 | 1.39 – 2.40 | < 0.0001 |
| Dialysis | 2.22 | 1.51 – 3.28 | < 0.0001 |
| Mechanical ventilation | 2.11 | 1.64 – 2.73 | < 0.0001 |

Abbreviations: HR, hazard ratio. CI, confidence interval.

### 3.3 Construction and Validation of Nomogram

Based on the results of multivariate analysis, the risk factors listed in Table 3 were used to construct a nomogram for 4-year OS (Figure 2), the prediction performance of which was further confirmed with C-index, ROC and calibration curve. The C-index for OS prediction in the training and validation set was 0.788 (95%CI: 0.763 to 0.813) and 0.783 (95%CI: 0.748 to 0.818), respectively. The AUC of ROC curve was 0.834 in training cohort and 0.832 in validation cohort (Figure 3). All these results above showed a great discriminatory ability of the nomogram in predicting the longterm OS of patients with AMI. Besides, the calibration curve indicates that there was a good consistency between the predicted and observed 4-year OS rates (Figure 4).

**Figure 2.**
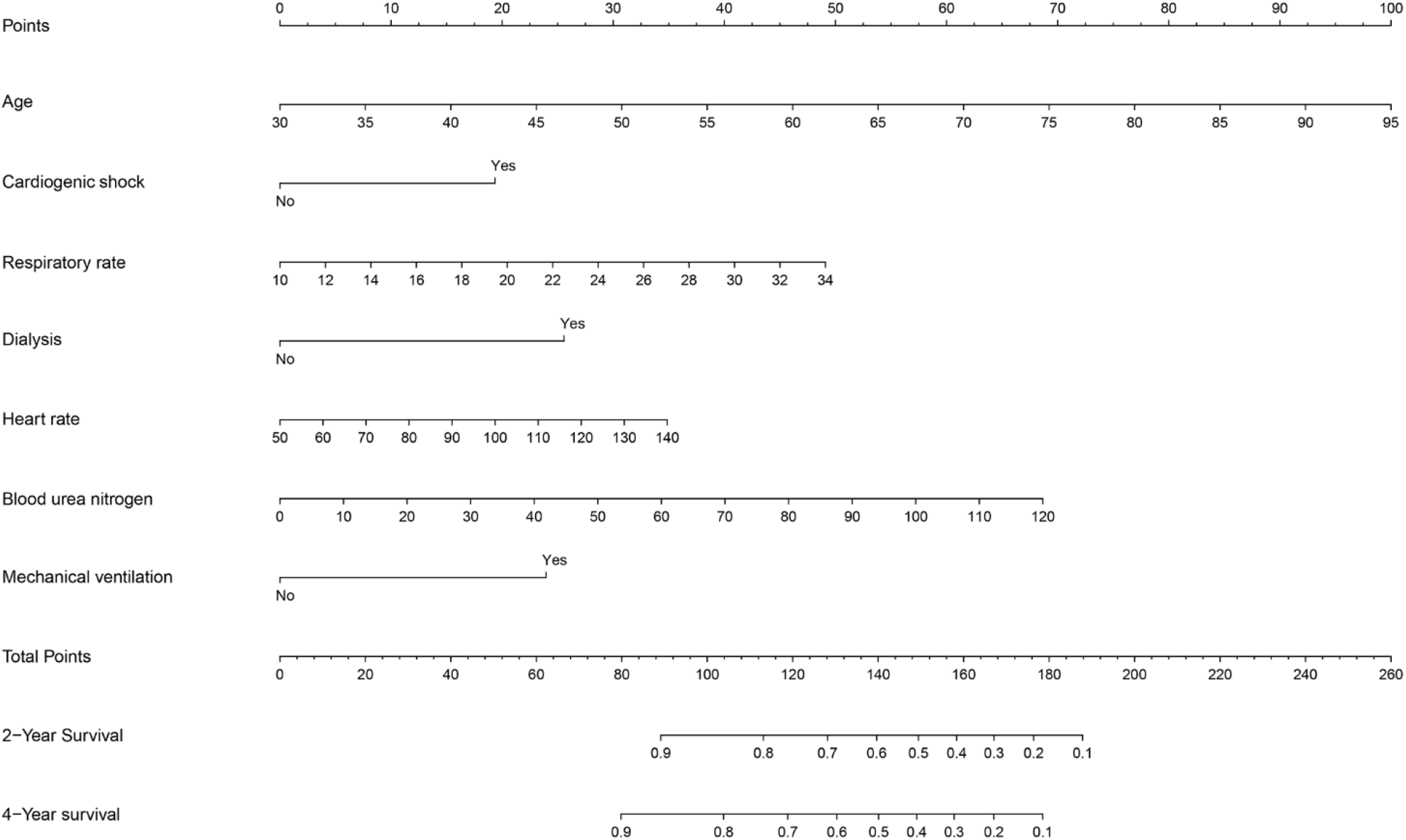
The nomogram for predicting 2- and 4-year overall survival of AMI. The nomogram included seven variables, including age, heart rate, respiratory rate, blood urea nitrogen, cardiogenic shock, dialysis, and mechanical ventilation. When use it, we should draw a vertical line upward from each variables to the “Points” line to get the score, and then add them up to get the total score. Last, draw a vertical line downward from the “Total Points” and get the 2- and 4-year survival of patients with AMI. AMI, acute myocardial infarction.

**Figure 3.**
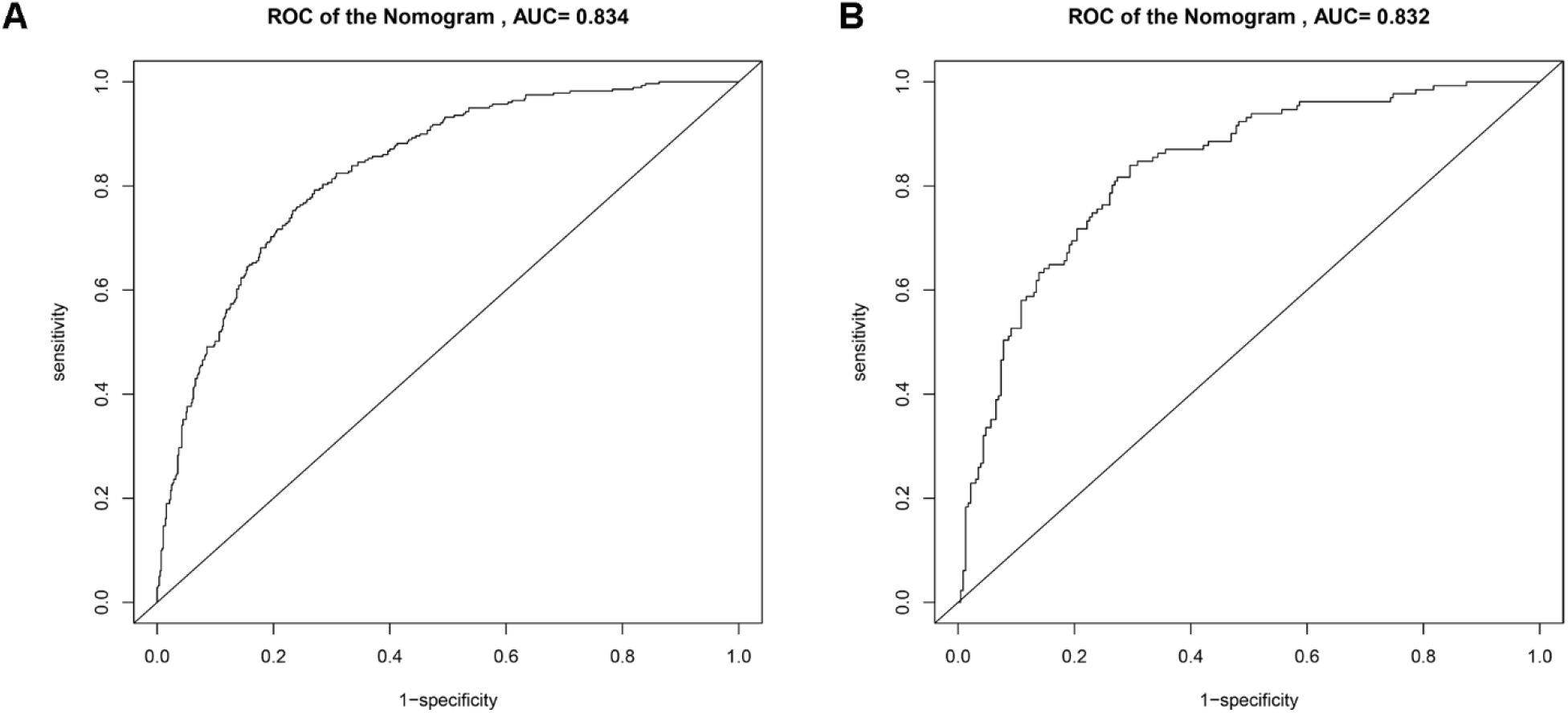
The ROC curve of the nomogram for predicting 4-year overall survival in training set (A) and validation set (B). The AUC of ROC curve in training and validation set was 0.834 and 0.832, respectively. X-axis: 1-specificity; Y-axis: sensitivity. ROC, receiver operating characteristic curve. AUC, area under the curve.

**Figure 4.**
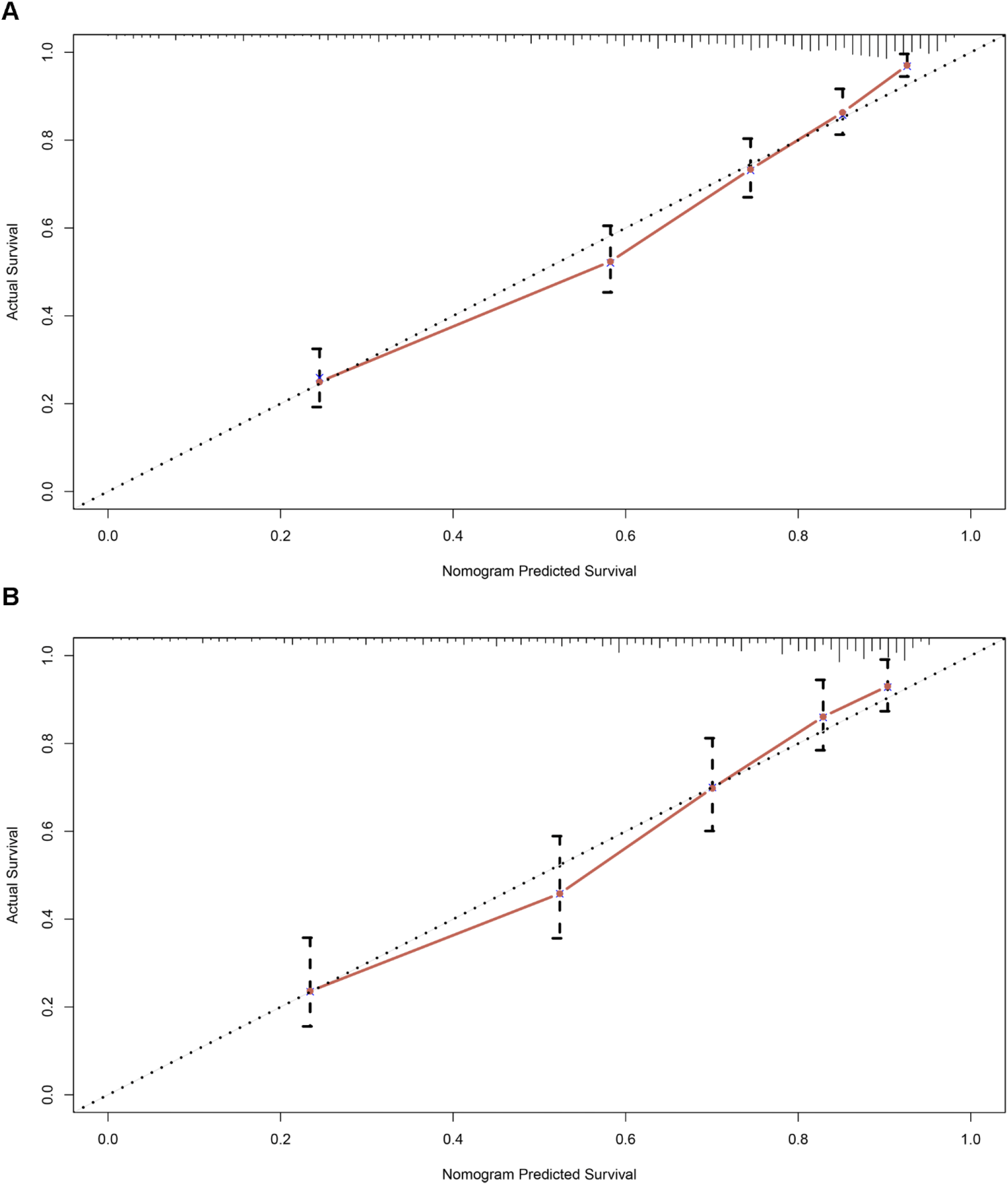
The calibration curve of the nomogram for predicting 4-year overall survival in training set (A) and validation set (B). The dotted line represents the ideal curve where the predicted value is the same as the observed value. X-axis: the predicted survival of the nomogram; Y-axis: the actual survival in the cohort.

## 4 Discussion

With the development of the ageing society, AMI has become one of the most common cause of death and disability worldwide [20]. Therefore, there is an urgent need for patients with AMI to more precise risk stratification methods, individual treatment and follow-up strategies. The nomogram is a visual medical prediction model that can provide accurate and personalized predictions for patient OS, and allows clinicians to make standardized clinical decisions [21].

In the present study, based on the large sample database MIMIC-III, a nomogram was developed and validated to predict the 4-year OS of patients with AMI by using these variables screened by univariate and multivariate analysis, including age, HR, RR, BUN, cardiogenic shock, dialysis, and mechanical ventilation. In previous studies, some nomograms have been reported for AMI, but they were mainly concerned with the short-term mortality of patients and the occurrence of complications, such as bleeding and acute kidney injury [22, 23]. To the best of our knowledge, this is the first time to establish a nomogram to predict the long-term survival of patients with AMI. In this prediction model, the predictors included had the characteristic of easy to obtain and calculate, including vital signs such as HR, and routine laboratory tests such as BUN, which have extremely high availability, especially in some economically underdeveloped areas. what’ more, the proposed nomogram has great discrimination and accuracy with a high C-index and well-done calibration curves, which can provide a reliable reference for clinical decision-making.

Through statistical analysis, it was found that the following seven factors were independently related to the long-term prognosis of AMI: age, HR, RR, BUN, cardiogenic shock, and the use of dialysis and mechanical ventilation, which was partly consistent with previous research. Compared with the young, the elderly are a unique population, often accompanied by structural and functional abnormalities of the heart, with a higher incidence of cardiovascular disease [24]. And age is also an independent predictor of the prognosis of patients with AMI. Chua et al. [25] has demonstrated that the short-term adverse events, including re-infarction, heart failure and mortality, increased significantly with the increasing of age in patients undergoing PCI for ST-elevation myocardial infarction (STEMI). Moreover, there was a higher middle- and long-term mortality for elderly patients with AMI [26, 27]. HR, as a very important vital sign, has been shown to be a risk factor for the poor prognosis of various cardiovascular diseases, including coronary artery disease [28]. Perne et al. [29] has explored that patients diagnosed AMI with a HR of 90 beats per minute or more at admission had a relatively low 3-month survival despite optimal treatment. In the STEMI patients undergoing primary PCI, for every 5 beats per minute increase in discharge HR, the 4-year cardiovascular mortality increased by 24% [30]. Both BUN and creatinine are the end-products of nitrogenous substances, and they are also the most commonly used indicators to reflect renal function in clinical practice [31]. In addition, BUN levels are also affected by the status of low cardiac output, insufficient systemic and renal perfusion, and activation of the neurohumoral system, which occur usually in the early stages of AMI. Some studies has shown that elevated BUN at admission is an significant marker for in-hospital and long-term mortality in patients with AMI, after adjusting for creatinine and other potential covariable [32, 33]. Respiratory failure and acute kidney injury are the common complications of AMI. Previous studies have indicated that about 8% of patients with AMI required mechanical ventilation, and 3-4% of patients were treated with hemodialysis or other forms of renal replacement therapy [34, 35]. And the use of these interventions has been proved to be significantly associated with the poor survival of patients [35, 36], which is the same as our results.

Our research also had some limitations as follows. First, the clinical data for analysis was extracted from a single-center institution, and the representativeness of samples was limited to some extent. Then, the vital signs, laboratory tests and other variables were primarily derived from the data of patients within 24 hours after ICU admission, which may cause a certain degree of selection bias. Third, the indicators included in our study were mainly conventional and easily accessible parameters, some specific indicators, such as N-terminal pro brain natriuretic peptide and echocardiography parameters, cannot be included due to the large number of missing values, which may reduce the accuracy of the model. Last, we did not conduct external validation of our model by using our own data.

## 5 Conclusion

The nomogram we established can effectively predict the 4-year OS of critical ill patients with AMI, and the results of validation shown that it has an accurate predictive performance and can provide a good reference for evaluating long-term survival of these patients.

## Data Availability

Publicly available datasets were analyzed in this study. This data can be extracted from Monitoring in
Intensive Care Database III version 1.4 (MIMIC-III v.1.4) after passing on the required courses and
obtaining the authorization.

https://mimic.physionet.org/

## 6 Author Contributions

Author Z. X. Y and L.H.Z designed the research. Y.Y.T performed the data analysis and drafted the manuscript. X.F.Z, Y.L.F, W.C.L, and Z.H.L analyzed the data and revised the manuscript.

## 7 Conflicts of Interest

All authors declare that there is no conflict of interest.

## 8 Acknowledgement

Our study was supported by the National Natural Science Foundation of China (81873416) and the National Science and Technology Major Project (2017ZX0930401405).

## 9 Data availability

Publicly available datasets were analyzed in this study. This data can be extracted from Monitoring in Intensive Care Database III version 1.4 (MIMIC-III v.1.4) after passing on the required courses and obtaining the authorization.

## Notes

### Competing Interest Statement

The authors have declared no competing interest.

### Clinical Trial

This study is a retrospective study, and trial ID is not required.

### Author Declarations

Since this project has been approved by the institutional review boards of Massachusetts Institute of Technology (Cambridge, MA, USA) and Beth Israel Deaconess Medical Center (Boston, MA, USA) and the identifying elements related to patient privacy have been removed from the database, our research does not need to provide the additional approval of ethics committee.

